# Diet shapes the gut microbiome: cross-sectional and longitudinal insights from the Human Phenotype Project

**DOI:** 10.1101/2025.10.12.25337559

**Authors:** Tomer Segev, Daniel Barak, Liron Zahavi, Anastasia Godneva, Michal Rein, David Krongauz, Hagai Rossman, Adina Weinberger, Eran Segal

## Abstract

Diet is a major environmental factor influencing the human gut microbiome. However, the effects of specific foods and dietary patterns on microbial composition, diversity, and function is not fully understood, limiting progress toward personalized dietary strategies. Leveraging 10,064 participants from the Human Phenotype Project with app-based diet logs and shotgun metagenomics, we predicted diet-microbiome associations at species-level resolution. Diet significantly predicted microbial diversity (Richness r=0.24, Shannon index r=0.22), the relative abundance of 664 of 724 species tested (91.5%, FDR<0.05), and 313 of 320 functional pathways (97.8%, FDR<0.05). Feature attribution identified distinct food-microbe links, including coffee with Lawsonibacter asaccharolyticus (r=0.41), yogurt with Streptococcus thermophilus (r=0.38), and milk with Bifidobacterium species (r=0.31-0.35). In parallel, broader dietary patterns, especially those related to the degree of food processing, emerged as predictors of microbial diversity and composition. Longitudinal analyses on test set data showed that models remained predictive over two and four years (R²=0.73-0.79), with significant associations between observed and predicted changes in 115 and 92 species, respectively. Finally, we developed an exploratory analysis for suggesting personalized dietary interventions with predicted microbiome shift effects that are associated with improvements in key clinical biomarkers such as triglycerides. Overall, our findings establish diet as a key modulator of microbiome composition, diversity, and function, and highlight its potential for guiding personalized interventions.

## Introduction

The gut microbiome consists of trillions of microbes residing in the human digestive system. These microbes play a crucial role in digesting food and releasing metabolites, which can be both beneficial and harmful, and even impact the immune system and brain function^1^. The composition of the gut microbiome is influenced by various factors, including genetics, living environment, co-habitation, pets, diet, age, and sex^2^. Notably, most microbiome variation is attributable to environmental factors rather than genetics^3,4^.

Diet is a key factor shaping the composition and function of the gut microbiome^5^. For instance, diets rich in diverse, plant-based fibers are linked to increased microbial diversity and the proliferation of beneficial, butyrate-producing bacteria^6–8^. In contrast, "Western" diets, characterized by high intake of processed foods, fats, and simple sugars, are linked to reduced microbial diversity and the promotion of pro-inflammatory taxa^9–13^. These effects arise because undigested dietary components serve as the main substrates for microbial metabolism. The type of substrates available - such as complex plant fibers versus simple sugars - creates ecological pressures that favor taxa able to exploit them, directly shaping community composition and function^14^.

Although broad dietary patterns such as Mediterranean, vegan, vegetarian, and omnivore are known to affect the gut microbiome^15–17^, the effect of them and other patterns remain to be defined further. In parallel, the roles of individual foods and nutrients in shaping specific taxa are still poorly understood. Many studies rely on food-frequency questionnaires with limited resolution, small sample sizes, or cross-sectional only designs, making it difficult to pinpoint which individual foods, nutrients, or dietary patterns most strongly affect aspects of the gut microbiome. Moreover, the persistence of diet-microbiome associations over time, and their potential for guiding personalized dietary strategies, remain poorly defined.

To address these gaps, we leveraged data from the Human Phenotype Project (HPP)^18^, a large prospective longitudinal cohort with over 14,000 deeply-phenotyped individuals, including shotgun metagenomic microbiome profiles, detailed app-based diet logs, and follow-up visits over 2-4 years. This combination of scale, temporal depth, and high-resolution diet and microbiome data provides a unique opportunity to map the diet-microbiome relationship with unprecedented precision.

In this study, using the HPP, we set out to define how individual foods, nutrients, and dietary patterns shape microbiome composition, diversity and functionality. We further asked whether diet-microbiome associations observed at baseline persist over time, and whether personalized dietary simulations can predict microbiome shifts to inform tailored nutritional strategies. By combining large-scale, high-resolution dietary records with metagenomic profiling, our work deepens understanding of diet-microbiome relationships and highlights their potential for guiding individualized dietary interventions.

## Results

### Study population and design

We analyzed data from 10,064 participants in the Human Phenotype Project (HPP) who had both metagenomic microbiome profiles and dietary logs collected via a mobile application (see Methods for exclusion criteria). The HPP includes over 14,000 deeply phenotyped Israeli adults, predominantly healthy, primarily of Ashkenazi Jewish ancestry, aged 40-75, with follow-up visits every two-years.

We split the data into 8,102 participants at baseline visit only, reserving 1,962 participants as test set data for further analyses. Of the test set data participants, 1,576 returned for a two-year visit, and 1,294 returned for a four-year visit.

We trained models to predict microbiome features from dietary data and used feature importance analysis to identify specific dietary components contributing to each prediction (Fig. 1a). Age and sex were included as covariates in all models. We used both linear regression and gradient-boosted decision trees for these predictions. Tree-based models consistently outperformed linear models, indicating non-linear diet-microbiome relationships (Extended Fig. 1). All downstream analyses therefore used results from tree-based models. To assess model significance, we performed a permutation test. We randomly reassigned microbiome profiles across participants to generate a null distribution of model performance. Dietary data were derived from app-based food logs and included 700 features, comprising individual foods consumed by at least 1% of participants, food categories, nutrient values, and summary statistics (mean and coefficient of variation) of dietary patterns (e.g., number of foods consumed per day or week) and adherence scores (e.g., Mediterranean diet).

**Figure 1:**
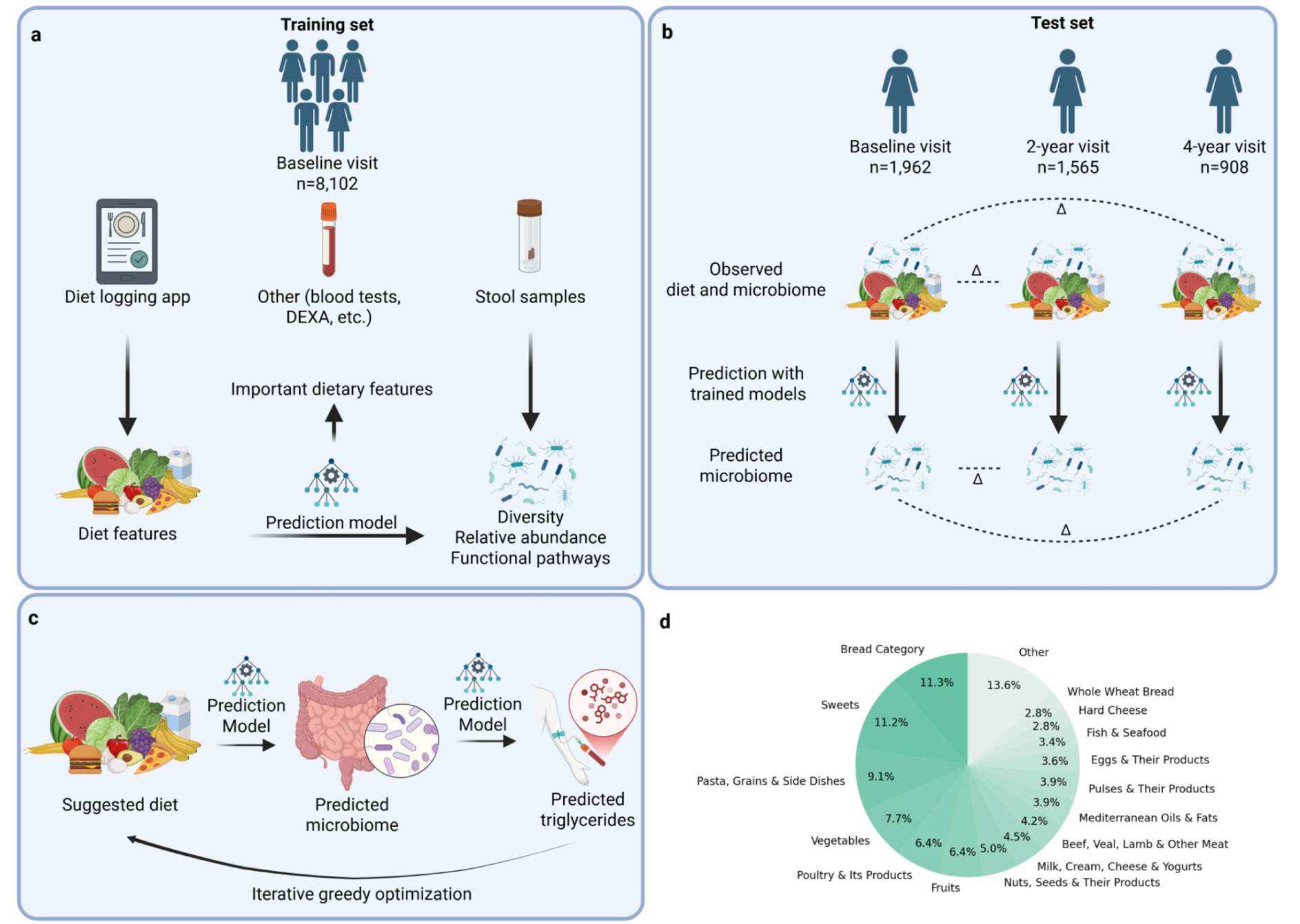
Overview of study design, longitudinal analysis, and personalized dietary simulations. **a.** Study design. We collected data from 10,064 participants in the Human Phenotype Project, including dietary logs from a mobile application, stool samples and other tests. We trained models on 8,102 baseline participants to predict microbiome diversity, taxonomic composition, and functional pathways from diet features, with age and sex included as covariates. **b.** Longitudinal analysis framework. In the test set, diet and microbiome were observed at baseline (n=1,962), two-year (n=1,565) and four-year (n=908) follow-ups. Models trained at baseline to predict microbiome composition from diet were used to estimate microbiome changes from dietary changes. Predicted changes were then correlated with observed changes. **c.** Personalized dietary simulation framework. We applied a two-stage modeling approach: first linking diet to the microbiome, then linking the microbiome to host phenotypes. An iterative greedy optimization algorithm generated individualized dietary modifications predicted to shift the microbiome toward profiles associated with improved phenotypes. We used triglycerides as an illustrative target due to their clinical relevance and strong predictability from microbiome features. **d.** Distribution of calorie intake across food categories in the cohort, showing mean percentages from baseline dietary records.

We included 13 diet adherence scores in our study: the Alternative Healthy Eating Index (AHEI)^19^, DASH score^20^, EAT-Lancet score^21^, healthful Plant-Based Diet Index (hPDI)^22^, Mediterranean diet adherence score (iMEDAS^23^), and calorie percentage based scores for whole-food plant-based, vegetarian, vegan, carnivore, and various low-carbohydrate diets. We also calculated the percentage of calories from ultra-processed foods (UPF percentage)^24^.

Microbiome targets included species-level taxonomic relative abundances, gut microbiome diversity metrics, and metabolic pathways derived from functional profiling.

For the microbiome-based health marker predictions, we used other body systems measured in the cohort, including blood biomarkers, body composition (e.g., DEXA-derived fat and muscle indices), cardiovascular measurements, and additional physiological traits.

### Gut microbiome diversity is predicted by diet

We investigated how dietary features contribute to gut microbial diversity, using both species richness and the Shannon index as measures of alpha diversity. Richness captures the number of distinct microbial species, while the Shannon index accounts for both the abundance and evenness of species in the community.

To examine this relationship, we trained gradient boosted decision trees using lightGBM (LGBM)^25^ to predict diversity metrics from dietary features. Our results show that dietary intake is a significant predictor of richness (r=0.24) and Shannon index (r=0.22), outperforming predictions from baseline models using only age and sex, and from models trained on permuted labels (Fig. 2a; see Methods). Sex alone explained a substantial fraction of the variance for Shannon diversity, consistent with previous findings that diversity is generally higher in females^26^.

**Figure 2:**
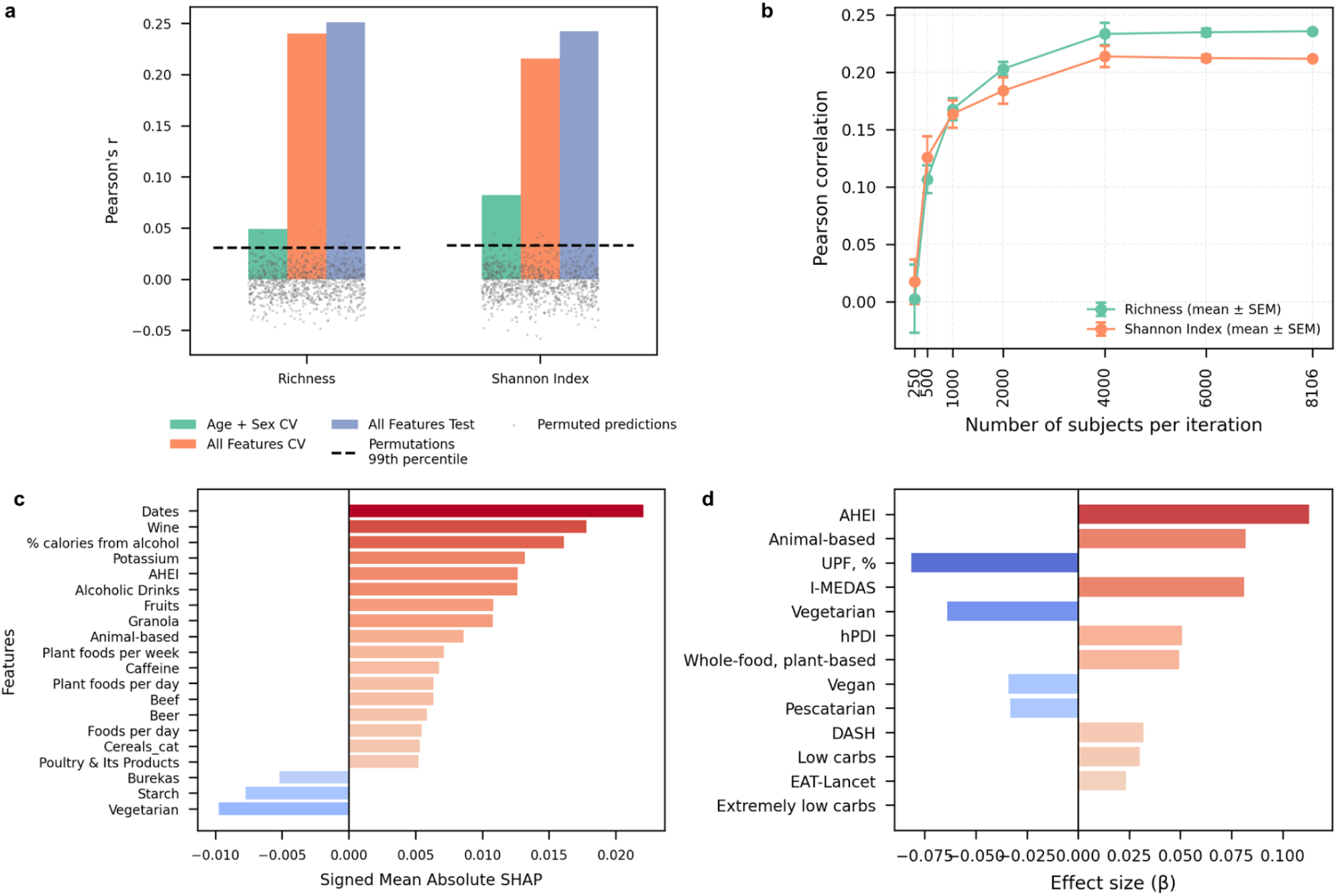
Dietary features predict gut microbial diversity. **a.** Pearson correlation coefficients (r) for LightGBM models trained to predict species richness and Shannon diversity using age and sex (blue) or full dietary data (orange). Grey dots represent permuted Pearson’s r with all features, and dashed line indicates 99th percentile of permutations.**b.** Model performance increases with sample size. Prediction improved sharply when scaling from hundreds to thousands of individuals, then began to plateau, based on five resampled subsets for each sample size. Both richness and Shannon index showed this trend. **c.** SHAP summary plot for species richness, shows feature contributions sorted by importance; plant-based and animal-derived whole foods, as well as wine intake, are contributors of higher richness, while processed items such as breads and pastries have negative impact. SHAP direction was derived from spearman correlation directions. **d.** Effect size (β) from a linear regression model predicting microbial richness and Shannon diversity from each diet score, adjusted for age and sex. Diets allowing for ultra-processed foods (UPF) showed negative effects. Red bars indicate a statistically significant association (q<0.05).

SHapley Additive exPlanations (SHAP)^27^ analysis revealed that both healthy and unhealthy dietary items strongly influenced the diversity measures (Fig. 2c). Consumption of unprocessed foods - both plant-based and animal-derived - was associated with increased richness. In contrast, ultra-processed foods and baked goods (e.g., burekas, white bread) were negatively associated with diversity. Notably, wine intake was positively linked with richness, and has been shown to increase microbial diversity and the abundance of beneficial bacteria such as Akkermansia in certain individuals^28^.

To assess how adherence to different dietary patterns affects the gut microbiome, we fitted linear regression models using diet adherence scores (see “Diet Adherence Scores” in Methods) to predict alpha diversity, adjusting for age and sex (Supplementary Table 3).

Food processing level emerged as the dominant dietary predictor of reduced microbial diversity (Fig. 2d). Among the tested scores, the Alternate Healthy Eating Index (AHEI), reflecting minimally processed, nutrient-rich intake, was the strongest positive predictor of microbial richness and Shannon diversity. In contrast, the UPF percentage, quantifying ultra-processed food intake, was the strongest negative predictor. Unexpectedly, the carnivore score also positively predicted microbial diversity - comparable to AHEI - despite encouraging absence of plant fiber. This finding challenges a simple plant-versus-animal food axis and suggests that avoiding ultra-processed foods may be as critical for maintaining gut diversity as the inclusion of specific food groups.

We also assessed the statistical power of our cohort by evaluating how predictive performance scales with sample size. Model accuracy rose sharply when moving from hundreds to thousands of individuals, then began to plateau. This pattern, observed across five resampled subsets for each sample size, indicates that our current cohort is large enough to capture the diet-diversity relationship, and further increases in size are unlikely to yield large gains (Fig. 2b).

### Species relative abundances are significantly predicted by diet

We trained models to predict species-level relative abundances from dietary features, controlling for age and sex. Out of 724 species analyzed, 664 (91.46%) yielded significant predictions (FDR < 0.05, Benjamini-Hochberg method^29^) in the lightGBM models (Fig. 3a), and 678 species (93.78%) were significant in the independent baseline test set (n = 1,962; Extended Fig. 1). To assess statistical power, we further examined how predictive performance scales with sample size. Prediction accuracy rose steeply when moving from hundreds to thousands of individuals and then plateaued, a pattern consistent across five resampled subsets at each sample size for the top 100 most predictive species (Fig. 3b).

**Figure 3:**
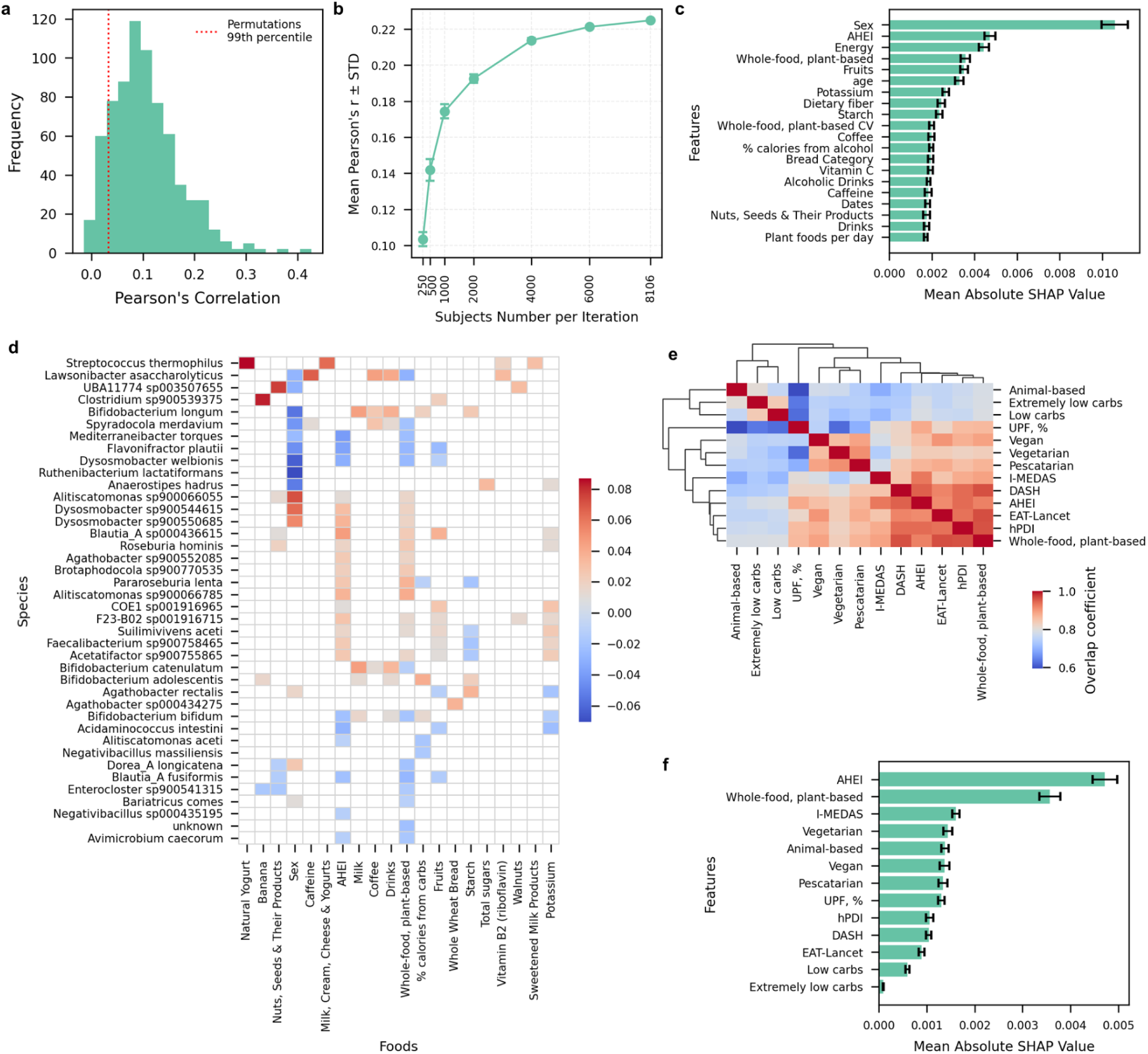
Dietary features predict gut taxonomic composition. **a.** Histogram of Pearson correlations (LGBM) between predicted and observed species abundances. A red line indicates the 99th percentile of correlations obtained from permutation testing, highlighting significant correlations. **b.** Model performance improves with sample size, shown by mean ± s.e.m. of Pearson’s r across the top 100 predicted species. Prediction rose steeply from hundreds to thousands of individuals and then plateaued, based on five resampled subsets per sample size. **c.** Mean absolute SHAP values averaged across species; error bars show SEM across species. Higher values indicate greater average importance for the model’s predictions. **d.** Main predictors heatmap of SHAP values for the top 40 most predictable species and the 20 dietary features with the highest maximal importance. We clustered species by similarity in their SHAP value profiles across foods. SHAP values < 0.01 were masked. **e.** Overlap coefficient heatmap of species significantly associated with each diet score. Hierarchical clustering revealed four dietary groups with shared microbiome signatures: (1) whole-food-based Mediterranean-like diets (IMEDAS, AHEI, EAT-Lancet, WFPB, hPDI); (2) plant-based diets permitting processed items (vegetarian, vegan, pescatarian); (3) the UPF percentage score alone; and (4) low-carbohydrate, animal-based diets (carnivore, low-carb, extremely low-carb). **f.** Mean absolute SHAP values averaged across species, showing only dietary pattern features; error bars show SEM across species. Higher values indicate greater average importance for the model’s predictions. Higher values indicate greater overall importance in species-level prediction models.

We then used SHAP to identify which dietary features most significantly contributed to microbial abundance predictions. Species with the strongest diet-based predictions included Lawsonibacter asaccharolyticus (r=0.41), a butyrate producing microbe, predicted by coffee consumption, consistent with previous research^30^; Streptococcus thermophilus (r=0.38), highly predicted by yogurt consumption and used for its fermentation; The probiotics^31^ Bifidobacterium longum (r=0.35) and Bifidobacterium adolescentis (r=0.31), predicted by milk^32,33^, dairy and bread consumption^34,35^; the species UBA11774 sp003507655 (r=0.38), which is predicted by nuts consumption, and previously shown to mediate the effect of oils and fat consumption on Hb1ac outcome^36^; and Clostridium sp900539375, highly predicted by banana consumption and other fruits to a lesser extent (Fig. 3d).

Despite being a butyrate producer, Lawsonibacter asaccharolyticus is predicted negatively by dietary fiber consumption. Prior findings suggest polyphenols in coffee drive its abundance^30^, so the negative fiber contribution may be due to competitive interactions among fiber-utilizing microbes.

To assess how adherence to different dietary patterns affects the gut microbiome, we fitted linear regression models using diet adherence scores (see “Diet Adherence Scores” in Methods) to predict species abundance, adjusting for age and sex (Supplementary Table 3). In addition to the effects of food processing on diversity, dietary patterns grouped into four major microbiome composition clusters at the species level (Fig. 3e): (1) whole-food-based Mediterranean-like diets (IMEDAS, AHEI, EAT-Lancet, WFPB, hPDI); (2) plant-based diets permitting processed items (vegetarian, vegan, pescatarian); (3) the UPF percentage score alone; and (4) low-carbohydrate, animal-based diets (carnivore, low-carb, extremely low-carb). Microbial composition differed across these clusters. For instance, Alistipes aceti and Negativibacillus species were enriched in the carnivore group and depleted in vegetarian and pescatarian diets, reflecting the influence of specific food choices.

Together, these results show that diet predicts the relative abundances of most gut microbial species, with predictions revealing specific food-microbe links, some of which align with known associations and others suggesting novel dietary influence on the microbiome.

### Food-derived microbial species

Using curated Food Metagenomic Data (cFMD^37^), we tested whether significantly predicted species occur naturally in foods and thus enter the gut upon consumption (Extended Fig. 3). Although coffee consumption strongly predicted Lawsonibacter asaccharolyticus (r=0.41), we did not detect it in coffee sample sequencing. As expected, Streptococcus thermophilus (r=0.38) was present in yogurt in high abundance, since it is used for yogurt fermentation. Milk, the top predictor for Bifidobacterium, contained several species, including B. longum (r=0.35) and B. adolescentis (r=0.31). Interestingly, Bifidobacterium was not detected in sourdough bread despite the model’s bread predictions.

Notably, UBA11774 sp003507655 (r=0.38), mainly predicted by nut consumption, was detected in dark chocolate - a secondary but strong predictor for this species. These findings indicate that several diet-predictive species are also present in corresponding foods, supporting direct dietary transfer for some taxa as a mechanism of influence. Other taxa may reflect indirect ecological effects or unexplored dietary effects.

### Diet predicts microbial biosynthesis and degradation pathways

We next asked whether the functional potential of the microbiome, beyond its taxonomic composition, could also be predicted from diet (Extended Figs. 4, 5). To profile microbial metabolic pathways, we used HUMAnN^38^ on the stool metagenomes. Our analysis revealed significant associations between dietary features and a range of microbial metabolic pathways. Out of 320 pathways, 313 had significant predictions (FDR < 0.05). The pathways most predicted by diet (predictive r gain from baseline > 0.1) included microbial biosynthesis; carbohydrate and polysaccharide degradation; and plant-derived pathways.

High-quality diets rich in unprocessed plant and whole foods strongly predicted increased microbial biosynthesis of key metabolites, including amino acids and purines (Extended Fig. 4). The aromatic amino acid superpathway (COMPLETE-ARO-PWY) correlated positively with AHEI scores, fruit intake, and specific items such as dates and beef. In contrast, it was negatively associated with processed carbohydrates like pita and burekas. Similarly, pathways for L-arginine (ARGSYN-PWY) and de novo purine biosynthesis (PWY-841) were positively linked to fruits (e.g., watermelon, banana), potassium, and overall dietary quality, but suppressed by refined carbohydrates and high UPF scores. A notable exception was the folate transformation pathway (1CMET2-PWY), enriched in diets high in saturated fats and cholesterol, and depleted in plant-based diets. These patterns suggest that unprocessed, plant-rich diets support a microbiome with enhanced biosynthetic capacity for essential compounds.

Diet-microbiome associations extended to carbohydrate degradation pathways (Extended Fig. 4). Lactose degradation (LACTOSECAT-PWY) was predicted almost exclusively by dairy intake, while breakdown of plant polysaccharides - such as stachyose (PWY-6527), D-galactose (PWY-6317), and uronic acids (e.g., GLUCUROCAT-PWY, GALACTUROCAT-PWY, PWY-7242) - was positively associated with fruit and vegetable consumption and high diet quality. Refined carbohydrate intake consistently showed a strong negative association, pointing to a dichotomy in microbial specialization for complex plant substrates versus simple starches.

Plant-derived photosynthesis pathway (PWY-101) was highly predicted with high consumption of seeds, fruits and vegetables, and dropped with increasing UPF percentages. This pathway might reflect the presence of plant-derived DNA in stool, rather than a functional activity of the microbiome. In addition, seleno-amino acid biosynthesis (PWY-6936) pathway was highly predicted by plant food consumption and fish - a source of dietary selenium^39^.

An examination of microbial butyrate production, known for its health benefits^40,41^, revealed that its core biosynthetic pathways were weak predictors within the model. Specifically, pathways such as acetyl-CoA fermentation to butanoate (PWY-5676) and pyruvate fermentation to butanoate (CENTFERM-PWY) exhibited low correlation values (r=0.09 and r=0.07, respectively). Despite this low predictive power, the qualitative dietary associations for these pathways aligned with established nutritional science. Higher predicted butyrate production was positively associated with the intake of fruits, vegetables, and salmon. Conversely, a negative association was observed with bread and rice consumption, a finding that may reflect the influence of refined grains, which have been shown to reduce butyrate levels - in contrast to the known positive effect of their whole-grain counterparts^35,42,43^.

### Diet-microbiome associations persist longitudinally

We next tested whether diet-microbiome associations observed at baseline persist over time (Fig. 1b). We applied models trained on baseline data (n=8,102) to test set participants to predict microbial relative abundances at baseline (n=1,962), two-year (n=1,565), and four-year (n=908) visits. These predictions reflect the relationship between diet and the microbiome, as captured by the cross-validation (Fig. 4a).

**Figure 4:**
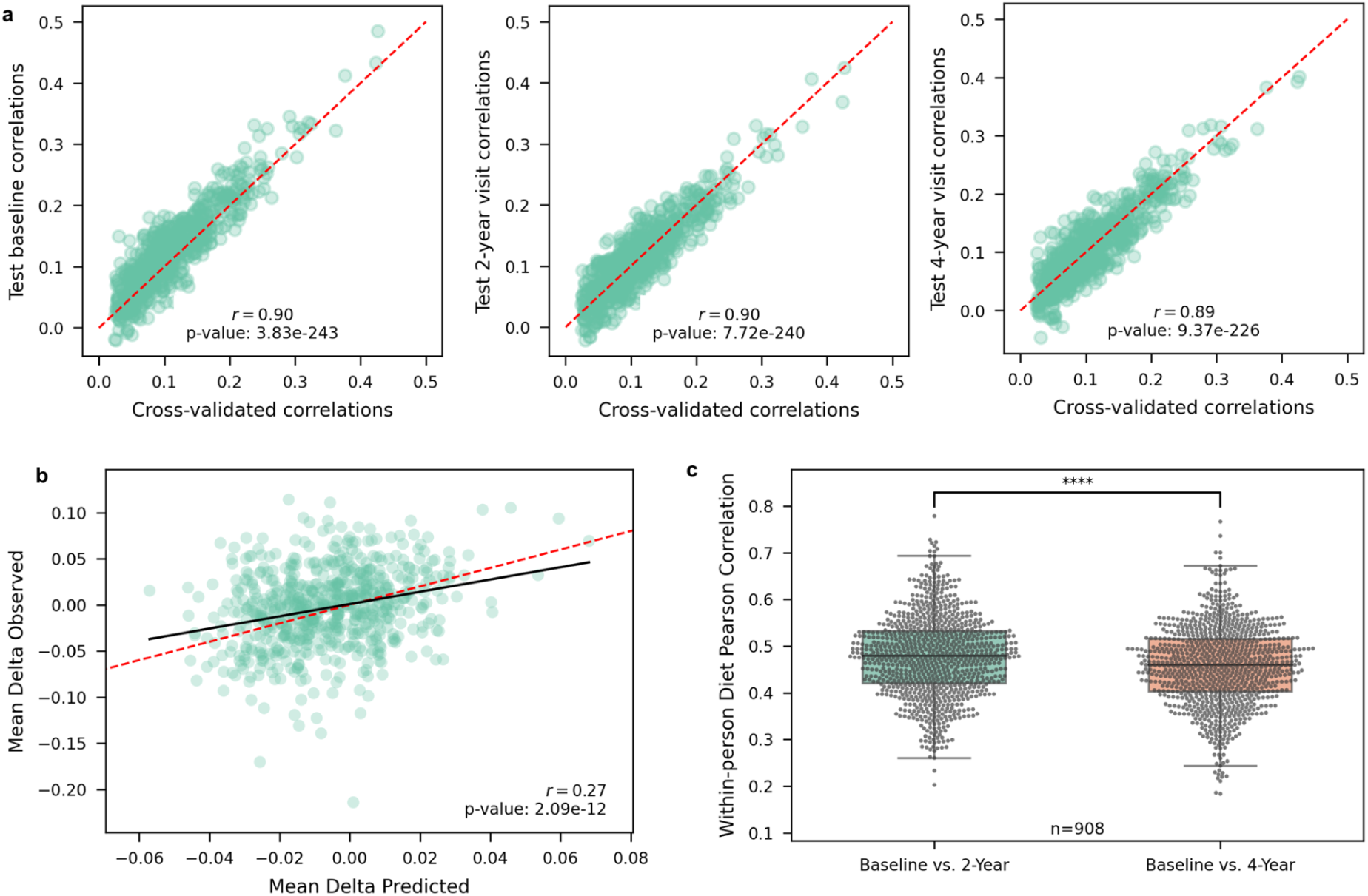
Diet-microbiome associations persist over two- and four-year follow-ups. **a.** Model performance at baseline and follow-ups. Scatter plots compare cross-validated predictions with test set predictions. Strong agreement was observed at all time points (baseline r=0.9; two-year visit r=0.9; four-year visit r=0.89). The red dashed line indicates the line of identity. **b.** Association of predicted and observed microbial changes. Each point represents a microbial species, with mean predicted change (baseline to two-year follow-up) plotted against mean observed change. Species predicted to increase tended to rise in abundance, whereas those predicted to decrease declined, supporting the validity of diet-microbiome links over time. **c.** Dietary self-consistency over time. Each dot indicates within-person Pearson correlation between baseline and follow-up dietary intake binarized vectors. Median correlation declined from two-year to four-year follow-up (Wilcoxon P < 1×10⁻⁴), reflecting dietary drift over time.

To assess whether diet changes are associated with relative abundance changes, we calculated the change in observed and predicted microbial abundances between the baseline and visit time points (Fig. 4b), and correlated these changes. Significant correlations indicate that changes in observed microbial abundances align with those predicted from dietary changes. Correlations were corrected for multiple testing using the Benjamini-Hochberg method (FDR < 0.05), but species were ranked by their cross-validation prediction strength.

Of the 664 species with significant baseline predictions, 115 showed significant correlations between observed and predicted changes at the two-year visit, and 92 at the four-year visit (Supplementary Table 4). These correlations included top-ranked taxa such as Lawsonibacter asaccharolyticus, Streptococcus thermophilus, Bifidobacterium species, and UBA11774 sp003507655. Species with higher baseline predictability were more likely to show significant longitudinal correlations (Supplementary Table 4).

The lower number of significant correlations at the four-year visit may be due to a smaller sample size. The number of significant correlations did not differ between visits when the two-year analysis was restricted to the same individuals available at four years (McNemar p>0.05), supporting the interpretation that the smaller four-year signal reflects reduced sample size.

### Personalized dietary simulation model for microbiome health

Given the established role of the gut microbiome in regulating host metabolism and immune function, and the predictive relationship shown in this study between diet and microbiome, we investigated how dietary changes can affect microbiome composition in ways predicted to improve clinically relevant health phenotypes. To define the microbiome composition related to these health markers, we trained LightGBM regression models to predict these phenotypes from the gut microbiome taxonomic profiles, and compared the predictive performance to a baseline model including only age and sex. All phenotypes shown could be significantly predicted by the full model (age, sex, and microbiome), based on an FDR < 0.05 threshold (Extended Fig. 6).

We adopted a two-stage design, first modeling how diet relates to the gut microbiome and then how the microbiome relates to health phenotypes (Fig. 1c). This reflects the biological understanding that dietary effects on host metabolism and immunity are often mediated through the microbiome rather than acting directly. By structuring the models in this way, we can propose diet changes that are predicted to shift the microbiome in directions associated with improved phenotypes. Similar multi-stage approaches have been used previously; for example, Larsen and Dai (2022) constructed a sequence of models linking diet, microbiome composition, microbial functions, and host phenotypes^44^. To implement this, we applied a greedy search algorithm that iteratively optimized the target phenotype for each individual by evaluating incremental dietary change. The algorithm is personalized, based on the individual’s current diet and the relative abundances of their existing microbiome. The optimization relied on the two pretrained models capturing diet-microbiome and microbiome-phenotype associations. The algorithm was then applied to the test set subset of 1,962 baseline participants, offering personalized dietary changes and predicting microbiome shifts alongside corresponding improvements in the previously mentioned health phenotypes.

For downstream dietary simulation modeling, triglycerides were selected as a representative microbiome-associated phenotype due to their clinical relevance, and strong predictability from gut microbiome features and its emerging links to metabolic disorders such as hyperlipidemia^45,46^. As additional examples, we also applied the simulation framework to total visceral adipose tissue (VAT) mass and white blood cell count (WBC), which were similarly well-predicted and clinically relevant (Extended Figs. 7, 8).

Using triglycerides as the representative health phenotype for optimization, we illustrate in an example of personalized dietary recommendations for a single individual (56-60 years old male; Fig. 5a) alongside the corresponding predicted microbial shifts (Fig. 5b). Across the cohort, the model identified food groups that were frequently recommended for modification (Fig. 5c), most commonly reductions in high-glycemic or lipid-elevating foods such as rice, dates, beef, and wine, and increases in fruits such as apples and bananas and unsaturated fat sources such as almonds, walnuts, avocado, and tahini. The microbial species shown in Fig. 5d were those most frequently suggested for modulation by the model. Most individuals received only a limited number of recommended dietary changes (median = 2) with minimal net changes in total caloric intake (Fig. 5e). Finally, the model predicted triglyceride reductions across all individuals, with most experiencing modest improvements and a subset showing larger decreases. The representative individual from panels a and b was predicted to decrease by 3.2 mg/dL (Fig. 5f).

**Figure 5:**
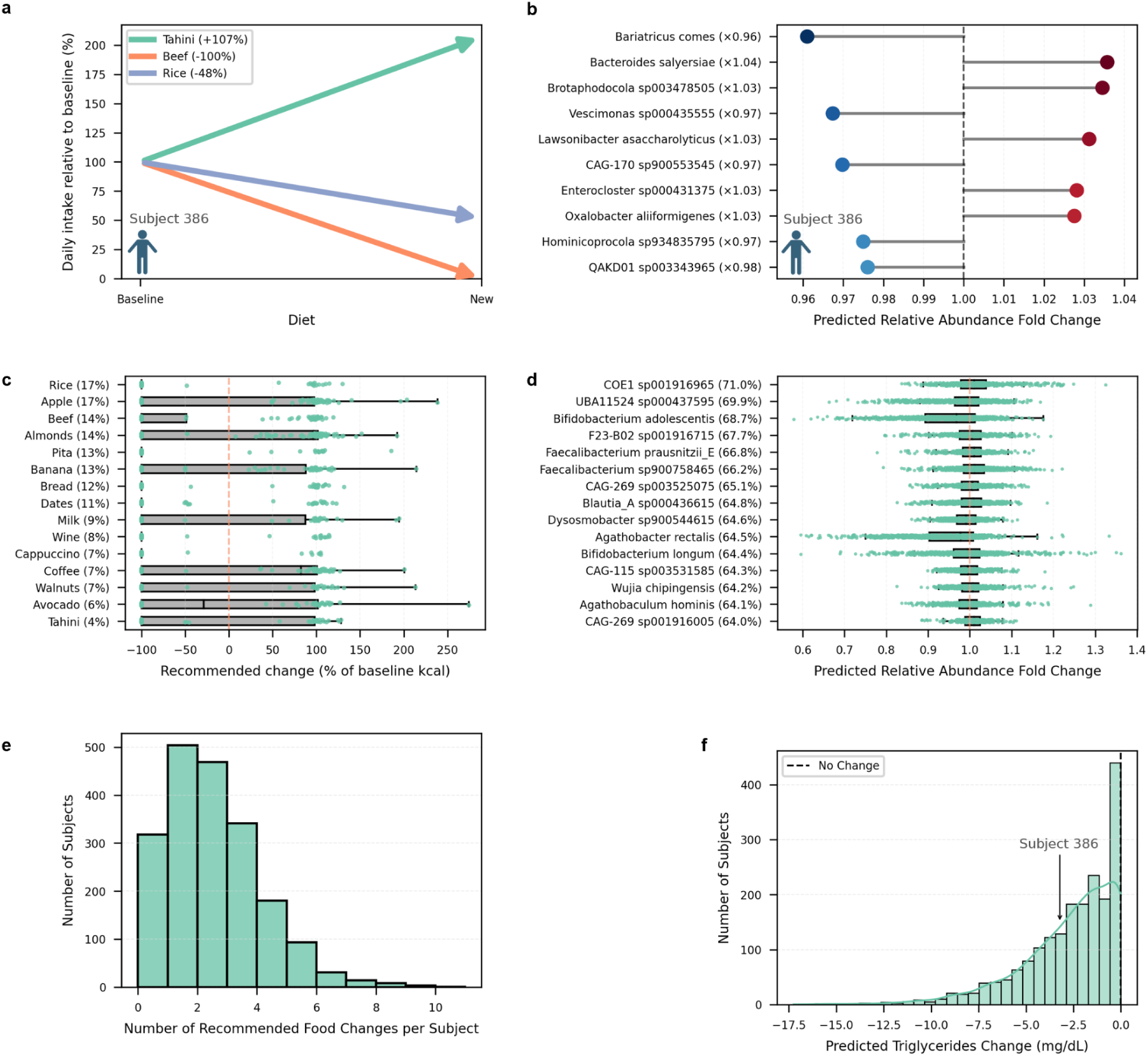
Personalized dietary simulated interventions predict microbiome shifts and triglyceride reductions. **a.** Personalized dietary recommendations for a representative individual (56-60 years old male). Arrows indicate foods predicted to increase or decrease relative to baseline intake, with percent changes shown in the legend. **b.** Predicted microbial shifts for the same individual, showing the ten species with the largest absolute fold changes. **c.** Most frequent food changes recommended across the cohort. Percentages show how many individuals who ate each food were advised to change it. Recommended changes are given as percent of baseline calories; each point is one individual. **d.** Microbial species most frequently predicted to change in response to the recommended diets. Percentages show how many individuals carrying each species were predicted to see a change. **e.** Number of recommended food changes per individual. Most people received only a few changes, with a median of two. **f.** Predicted changes in triglyceride levels across individuals. The dashed line marks no change, and the arrow highlights a predicted 3.2 mg/dL decrease in blood triglyceride concentration for the representative individual shown in panels a and b.

## Discussion

Using a cohort of 10,064 individuals from the Human Phenotype Project, we found that dietary intake significantly predicts alpha diversity, microbial pathways and species abundance for the majority of analyzed microbes. Feature importance analysis highlighted that specific dietary items, such as coffee, yogurt, dairy, nuts and certain fruits, strongly predict the abundance of key species including Lawsonibacter asaccharolyticus, Streptococcus thermophilus, and Bifidobacterium species. In addition, broad dietary patterns influenced microbiome composition and diversity, with the degree of food processing emerging as a primary driver. The mechanisms linking processed food to reduced microbial diversity remain unclear, but emerging evidence suggests that emulsifiers may contribute to this effect^47,48^. The predicted associations persisted over time, as shown in our longitudinal analysis. Based on these findings, we proposed a preliminary framework for personalized dietary interventions targeting microbiome health, which warrants future validation.

Our findings are consistent with other large-scale cohorts, who reported similar associations^49,15^. Compared to prior work, our study offers several advantages. First, its large sample size enabled sufficient power to detect robust associations, including at the species level. Second, our longitudinal design allowed us to assess whether baseline associations with diet persist over time. Third, the use of app-based dietary logging provided high-resolution, real-world intake data across thousands of participants. Together, these features strengthen the reliability of our findings.

However, several limitations should be considered. First, dietary intake was self-reported, which introduces potential biases from underreporting, recall error, or selective logging - even when entries were recorded via a smartphone app. While digital tools improve accuracy relative to retrospective questionnaires, they remain imperfect proxies for actual intake. Second, dietary data were collected over a 16-day period surrounding stool sampling.

Although this window likely reflects habitual intake for most individuals, it may miss longer-term patterns or recent dietary shifts that influence the microbiome. Third, the study population reflects dietary habits and microbial compositions typical of Israeli adults aged 40-70, which limits generalizability to other ages, disease states, and settings with different diets, lifestyles, or exposures. For instance, despite previously reported benefits of fermented foods on the microbiome^50–52^, we did not detect significant associations in our cohort, possibly due to low consumption of fermented foods other than yogurt. Fourth, although our models adjusted for age and sex, residual confounding is possible: sensitivity analyses that adjust for additional covariates such as BMI, medications and supplements, smoking, physical activity, recent antibiotics or probiotics, and stool consistency could help assess robustness. Finally, because interpretations rely on SHAP, collinearity among dietary features may diffuse attribution across correlated items; we therefore interpret feature-level attributions cautiously.

We developed a personalized dietary simulation model that identifies food changes predicted by our models to improve clinically relevant health markers through microbiome mediation. Using triglycerides as a representative target, the model simulated modest, individualized dietary changes, typically involving reductions in high-glycemic or lipid-elevating foods and increases in fiber-rich or unsaturated fat sources.

Among the most frequent recommendations were decreases in refined grains such as white rice, bread, and pita- high glycemic index foods known to raise postprandial triglycerides and recognized in clinical guidelines as targets for reduction in hypertriglyceridemia management^53,54^. Conversely, the model often recommended increasing fiber-rich fruits (e.g., apples, bananas) and unsaturated fat sources (e.g., almonds, walnuts, avocado, tahini) - a pattern consistent with dietary strategies known to reduce triglycerides, including the Mediterranean diet and macronutrient redistribution^55^.

The model predicted triglyceride reductions for all individuals, with most experiencing modest improvements and a subset showing larger decreases. These more responsive cases may represent microbiome or dietary profiles particularly receptive to targeted interventions. This in silico simulation is predictive rather than causal. While intriguing, these predictions remain model-based and require experimental validation in controlled dietary intervention studies to confirm both causal benefits and microbiome-mediated effects.

Overall, our findings strengthen our understanding of the relationship between diet and microbiome, and underscore the potential for diet-based modulation of the microbiome at species-level resolution. This paves the way for personalized dietary interventions to support gut health.

## Methods

### Ethics approval

Upon arrival at the study site, all participants were provided written informed consent. Personal identifiers were removed before any computational analyses were performed. The HPP cohort study complies with the principles of the Declaration of Helsinki and received approval from the Institutional Review Board (IRB) of the Weizmann Institute of Science (approval number 2392-4).

### Stool sampling, sequencing and taxonomic data processing

Stool collection, sequencing, and microbial profiling were performed as previously described^56^. Only microbial species that were identified in at least 5% of the samples were included, resulting in 724 species. Each stool sample was TSS normalized to 1, and log10-transformed relative abundances were used. Stool samples from two-year or four-year visits used the same species as baseline. Richness and Shannon index were calculated as features of alpha diversity.

### Dietary data processing

Dietary intake was recorded using a mobile application developed for the Human Phenotype Project (HPP). Entries were collected from two days before to 14 days after the baseline sampling. Entries missing names or energy values were excluded. Foods consumed by less than 1% of participants were removed, yielding 650 items. Foods with incomplete or incorrect nutrient data (except sugar substitutes), entries with implausible energy (<0 or >3000 kcal), or implausible weights (<0 or >2400 g) were excluded. Duplicate entries and days with total intake <500 or >4000 kcal, or 2.5 standard deviations below an individual’s mean intake, were also filtered out. Participants with fewer than three documented days were excluded. After filtering, 645 unique foods remained. Mean daily caloric intake was calculated for each item.

Nutrient data were added per food item. Macronutrient intake was expressed as mean daily calories; micronutrients were normalized by daily calorie intake. Additional features included fatty acid ratios. Eating behavior features (e.g., number of meals, unique foods per day, fasting hours, plant food diversity and ratios) were derived. Coefficient of variation across days was calculated for selected features. Thirty-three food categories were used to compute the mean daily percentage of caloric intake per category. Other features included total energy per day and the ratio of energy intake to estimated BMR. Dietary features from two-year or four-year visits used the same features as baseline.

### Diet adherence scores and patterns

Five standard diet scores were computed: IMEDAS: Adapted Mediterranean score emphasizing whole, minimally processed plant-based foods and traditional Israeli ingredients; EAT-Lancet: Plant-focused score based on planetary health principles; DASH: A heart-healthy diet pattern rich in fruits, vegetables, whole grains, and low-fat dairy; AHEI: A health-focused index emphasizing foods linked to reduced chronic disease risk; hPDI: Prioritizing healthy plant-based foods while penalizing unhealthy plant and all animal foods. Additional adherence scores were computed using predefined food categories. For each day, the score represented the percentage of calories from diet-aligned foods (range 0-1). Two penalties were incorporated: Base foods: Required minimum intake from core food groups. shortfalls incurred penalties. Moderation foods: Allowed only limited intake from certain categories; excesses incurred penalties.

The following adherence scores were derived: Whole Foods Plant-Based: Unprocessed plant foods only; Vegan: Includes processed vegan items; Vegetarian: Includes dairy, eggs and processed vegetarian items; Pescatarian: Vegetarian plus fish and seafood; Carnivore: Exclusively animal-based foods.

Other diet patterns included the percent of calories from NOVA 4 (ultra processed) foods, and the mean proportion of days classified as extremely low-carb (<10% carbs) or low-carb (10-26% carbs), based on daily carb intake thresholds.

Daily scores were aggregated into mean and standard deviation per individual. Scoring criteria are detailed in Supplementary Table 1. Diet classifications and NOVA scores of specific food items are detailed in Supplementary Table 2.

### Data split and standardization

The full dataset was split into training/validation (80%) and test (20%) subsets to enable unbiased performance evaluation. All non-binary input features (e.g., dietary variables) and microbial abundance targets were standardized using Z-score normalization. To avoid data leakage, the scaler was fit only on the training set, and the same parameters were applied to both training and test sets.

### Model training and hyperparameters

Separate models were trained for each microbial species and diversity metric using two approaches: Ridge linear regression and LightGBM. All training and hyperparameter tuning were performed within the training/validation set using 5-fold cross-validation.

Ridge Linear Regression: Ridge models were implemented using RidgeCV from scikit-learn^57^, which selects the optimal L2 regularization parameter (alpha) via internal cross-validation. We tested alphas=[0.01, 0.1, 0.5, 1, 10, 100].

LightGBM: Gradient-boosted trees were trained using the LightGBM regressor with the following fixed hyperparameters for regularization and efficiency: max_depth=4, n_estimators=2000, subsample=0.5, subsample_freq=1, colsample_bytree=0.3, learning_rate=0.001, n_jobs=8, random_state=1, reg_alpha=1, reg_lambda=1, min_data_in_leaf=20, verbosity=-1. For baseline models using only age and sex, colsample_bytree was set to 1.0 to prevent subsampling from two features.

### Performance Evaluation

Model performance was assessed using Pearson correlation (r) between predicted and true values. Out-of-fold predictions from cross-validation were concatenated to form a complete prediction vector (y’) for the training set, which was then correlated with the corresponding concatenated observed values (y). All modeling was performed in Python using scikit-learn. Final models were saved for use in held-out test prediction and longitudinal analysis.

### Permutation tests

Permutation tests were performed to assess the significance of cross-validated correlations. Species were stratified into 20 prevalence bins (0-5%, 5-10%, …, 95-100%). Microbiome targets were randomly reassigned to dietary data by sampling without replacement (seeds 1-1000), generating datasets in which each participant was paired with a random microbiome. After 1000 permutations, a prediction was considered significant (p < 0.001) if the correlation in the true data exceeded all correlations from permuted datasets. P values were corrected for multiple testing using the Benjamini-Hochberg method (FDR < 0.05).

### Sample size analysis

Sample-size scaling analyses were performed to evaluate statistical power. For each analysis, five independent subsets were drawn at each sample size by random sampling without replacement from the full training set. Within each subset, model accuracy was evaluated using 5-fold cross-validation. LightGBM models were trained for both alpha-diversity metrics and species-level abundances. For species-level analyses, the top 100 most predictive species were selected based on cross-validation performance. Model performance was computed as Pearson correlation (r) between predicted and observed values (Performance Evaluation; Methods). Mean performance and standard error across the five replicates were reported for each sample size.

### Feature importance analyses

To assess feature importance, models were retrained on the full training set without cross-validation. SHAP values were computed using TreeExplainer for LightGBM and LinearExplainer for Ridge. For each feature-target pair, mean absolute SHAP values were calculated. Targets with non-significant predictions were excluded. Analyses were based on the resulting feature-by-target matrix.

Mean absolute SHAP values averaged across species were computed for different feature groups, including all features (Fig. 3c) and diet adherence scores (Fig. 3f). Error bars show SEM across species. The direction of the mean absolute SHAP values were calculated by multiplying them with the sign of the Spearman correlation between the diet feature and microbiome target. Additional mean absolute SHAP values were averaged across microbial pathways, with each dot representing the SHAP value for an individual pathway (Extended Fig. 4). The main predictors heatmap was constructed using the best-predicted species and the features with the highest maximum mean SHAP values. Species were clustered by similarity in their SHAP value profiles across foods. Values < 0.01 were masked (Extended Fig. 5).

### Diet group comparisons

To identify differentially abundant species and assess the effect of diet patterns on microbial diversity, linear regression models were fitted to predict species abundance or diversity (y), from age (x₁), sex (x₂), and diet adherence score (x₃) as predictors. An intercept was included. The coefficient for x₃ was tested for statistical significance and adjusted using the Benjamini-Hochberg method (FDR<0.05). Since diet adherence scores were Z-score standardized, coefficients were directly comparable across diets.

To identify diets with similar microbial signatures, pairwise overlap coefficients were computed based on sets of significantly associated species, and hierarchical clustering was applied to group diets with similar profiles.

### Species presence in food items

The curated Food Metagenomic Database (cFMD), which contains metagenomes sampled from various foods, was used to identify microbial species derived from food consumption. Models were trained using MetaPhlAn^58^-derived species to match the taxonomic format used in cFMD. cFMD studies and microbial species were filtered to include only those with significant predictive associations and food items reported in the cohort. This filtering resulted in 153 species across 105 studies, corresponding to 36 food features. Further filtering was applied based on species presence, defined as abundance > 0, to retain only foods with at least one present species and species detected in at least one food. A heatmap was generated to visualize the final set of 16 foods and 38 species (Extended Fig. 3).

### Functional Pathways

Gut microbiome functional pathways were extracted from gut microbiome metagenomic data using HUMAnN3. These pathways were used as prediction targets in new models. Model training and feature importance analysis were performed using the same pipeline previously applied to taxonomic data.

### Longitudinal analyses

Longitudinal analyses were performed using the test cohort (n=1,962). Models trained on the baseline training set (n=8,102) were applied to predict microbial relative abundances at both baseline and the two-year follow-up. Changes in microbiome composition were computed by subtracting baseline values from those at follow-up, for both observed and predicted data. These changes were then correlated to assess the extent to which dietary changes predicted shifts in microbial abundances. Statistical significance was determined using the Benjamini-Hochberg method (FDR < 0.05), but correlations were ranked by the strength of cross-validated predictions, not by p-value. To enable direct comparison, the two-year analysis was also repeated in the subset of individuals with available four-year data, and significance overlap was evaluated using McNemar’s test.

### Health markers data processing

A dataset was constructed using measurements from each participant’s first (baseline) visit, integrating gut microbiome profiles with clinically relevant health phenotypes. These included blood biomarkers, body composition metrics derived from whole-body DEXA scans (e.g., fat mass index, ALM), cardiovascular measurements (e.g., blood pressure), and additional physiological traits. Total visceral adipose tissue (VAT) mass was obtained from DEXA scans. Fasting venous blood samples were used to measure triglycerides and other metabolic biomarkers, while white blood cell (WBC) counts were measured from the same samples but were not fasting-dependent. Each blood test was matched to the temporally closest stool sample within ±6 months. Gut microbiome features were taken from the processed stool data described above. For the present analysis, species with prevalence in >5% of participants and relative abundance >10⁻⁴ were retained, log₁₀ transformation was applied, and values were standardized to z-scores. Age and sex were retained as covariates, and outliers beyond ±1.5×IQR were excluded for each phenotype.

### Health markers prediction

Using the integrated dataset, gradient boosting regression models (LightGBM) were trained to predict WBC counts, total visceral adipose tissue (VAT) mass, and fasting triglycerides. For each phenotype, a baseline model including age and sex was compared with a microbiome-informed model that additionally incorporated species-level microbial abundances. Five-fold cross-validation was used to generate out-of-fold predictions. Predictive performance was assessed using Pearson correlation between observed and predicted values. P-values were adjusted for multiple testing, and only phenotypes with FDR < 0.05 were retained for downstream analyses.

### Diet simulation modeling

A personalized diet simulation model was developed to identify dietary modifications predicted to improve microbiome-mediated markers of metabolic and immune health, including VAT mass, triglycerides, and WBC counts. For each participant, baseline dietary intake and microbiome composition were used to simulate the effect of dietary changes. Predictions were generated using pre-trained LightGBM models linking diet to microbial abundances and microbial abundances to each target phenotype. A greedy search algorithm iteratively adjusted the intake of foods already present in each participant’s diet, first in 100% steps and, if no improvement was observed, in 50% steps. Candidate diets were constrained to the 5th-95th percentile of the cohort distribution, with a maximum ±20% change in daily calories; alcohol intake was limited to ≤5% of calories for women and ≤7% for men; and ultra-processed foods (NOVA 4) were not allowed to increase. Predicted microbial abundances were inverse-transformed from z-scores to estimated absolute abundances, and re-normalized to ensure proper compositionality, and used to estimate the target phenotype. For each participant, recommended dietary changes, predicted microbiome shifts, and estimated changes in VAT mass, triglycerides, and WBC counts were returned by the model.

## Data availability

The data used in this paper is part of the Human Phenotype Project (HPP) and is accessible to researchers from universities and other research institutions at: https://humanphenotypeproject.org/data-access.

Interested bona fide researchers should contact to obtain instructions for accessing the data

## Code availability

Code used in this study is available at the following GitHub link: https://github.com/TmrSegev/diet-microbiome

## Data Availability

The data used in this paper is part of the Human Phenotype Project (HPP) and is accessible to researchers from universities and other research institutions at: https://humanphenotypeproject.org/data-access.
Interested bona fide researchers should contact to obtain instructions for accessing the data

https://humanphenotypeproject.org/data-access

## Acknowledgements

We thank the members of the Segal group for fruitful discussions. E.S. is supported by the Crown Human Genome Center; the Larson Charitable Foundation New Scientist Fund; the Else Kröner Fresenius Foundation; the White Rose International Foundation; the Ben B. and Joyce E. Eisenberg Foundation; the Nissenbaum Family; Marcos Pinheiro de Andrade and Vanessa Buchheim; Lady Michelle Michels; Aliza Moussaieff; and grants funded by the Minerva Foundation, with funding from the Federal German Ministry for Education and Research and by the European Research Council and the Israel Science Foundation.

## Author contribution

T.S. conceived the project, designed and conducted all analyses, interpreted the results, and wrote the paper. D.B. designed and conducted the simulated dietary intervention and microbiome-phenotype prediction analyses, and wrote the paper. L.Z. and H.R guided the project and advised analyses. A.G. provided consultation and support for data processing. M.R. and D.K. advised diet adherence score development.

A.W. developed protocols, and oversaw sample collection and processing. E.S. conceived and directed the project and analyses, designed the analyses, interpreted the results, and wrote the paper.

## Extended Data Figures

Extended Data Fig. 1: Extended diversity and abundance predictions.

Extended Data Fig. 2: SHAP summary plots of highly predicted microbial species. Extended Data Fig. 3: Associations of food presence with food-derived microbial species in the cFMD dataset.

Extended Data Fig. 4: Microbiome functional pathways are highly predicted by diet. Extended Data Fig. 5: Main predictors heatmap of SHAP values for the most food-predictable pathways.

Extended Data Fig. 6: Predictive gain from baseline of various health phenotypes from gut microbiome.

Extended Data Fig. 7: Personalized dietary simulated interventions predict microbiome shifts and VAT reductions.

Extended Data Fig. 8: Personalized dietary simulated interventions predict microbiome shifts and WBC reductions.

## Supplementary Material

Supplementary Code Supplementary Tables 1-4:

Supplementary Table 1. Diet adherence scores calculation. Table 2. Foods diet classification and NOVA score. Table 3. Linear regression models for diet pattern comparisons. Table 4. Longitudinal analysis.

## References

1. Sanz, Y. et al. The gut microbiome connects nutrition and human health. Nat. Rev. Gastroenterol. Hepatol. 22, 534–555 (2025).

2. Kim, Y. S., Unno, T., Kim, B.-Y. & Park, M.-S. Sex Differences in Gut Microbiota. World J. Mens Health 38, 48–60 (2020).

3. Rothschild, D. et al. Environment dominates over host genetics in shaping human gut microbiota. Nature 555, 210–215 (2018).

4. Gacesa, R. et al. Environmental factors shaping the gut microbiome in a Dutch population. Nature 604, 732–739 (2022).

5. Carmody, R. N., Varady, K. & Turnbaugh, P. J. Digesting the complex metabolic effects of diet on the host and microbiome. Cell 187, 3857–3876 (2024).

6. Hills, R. D. et al. Gut Microbiome: Profound Implications for Diet and Disease. Nutrients 11, 1613 (2019).

7. Arora, T. et al. Microbial fermentation of flaxseed fibers modulates the transcriptome of GPR41-expressing enteroendocrine cells and protects mice against diet-induced obesity. Am. J. Physiol.-Endocrinol. Metab. 316, E453–E463 (2019).

8. McDonald, D. et al. American Gut: an Open Platform for Citizen Science Microbiome Research. mSystems 3, 10.1128/msystems.00031-18 (2018).

9. Simpson, H. L. & Campbell, B. J. Review article: dietary fibre–microbiota interactions. Aliment. Pharmacol. Ther. 42, 158–179 (2015).

10. Dostal Webster, A. et al. Influence of short-term changes in dietary sulfur on the relative abundances of intestinal sulfate-reducing bacteria. Gut Microbes 10, 447–457 (2019).

11. Malesza, I. J. et al. High-Fat, Western-Style Diet, Systemic Inflammation, and Gut Microbiota: A Narrative Review. Cells 10, 3164 (2021).

12. Rinninella, E. et al. The role of diet in shaping human gut microbiota. Best Pract. Res. Clin. Gastroenterol. 62–63, 101828 (2023).

13. Vangay, P. et al. US Immigration Westernizes the Human Gut Microbiome. Cell 175, 962–972.e10 (2018).

14. Sonnenburg, J. L. & Bäckhed, F. Diet–microbiota interactions as moderators of human metabolism. Nature 535, 56–64 (2016).

15. Fackelmann, G. et al. Gut microbiome signatures of vegan, vegetarian and omnivore diets and associated health outcomes across 21,561 individuals. Nat. Microbiol. 10, 41–52 (2025).

16. Gundogdu, A. & Nalbantoglu, O. U. The role of the Mediterranean diet in modulating the gut microbiome: A review of current evidence. Nutrition 114, 112118 (2023).

17. Ross, F. C. et al. The interplay between diet and the gut microbiome: implications for health and disease. Nat. Rev. Microbiol. 22, 671–686 (2024).

18. Reicher, L. et al. Deep phenotyping of health–disease continuum in the Human Phenotype Project. Nat. Med. 1–13 (2025) doi:10.1038/s41591-025-03790-9.

19. Chiuve, S. E. et al. Alternative Dietary Indices Both Strongly Predict Risk of Chronic Disease, ,. J. Nutr. 142, 1009–1018 (2012).

20. Karanja, N. M. et al. Descriptive Characteristics of the Dietary Patterns Used in the Dietary Approaches to Stop Hypertension Trial. J. Am. Diet. Assoc. 99, S19–S27 (1999).

21. Stubbendorff, A. et al. A systematic evaluation of seven different scores representing the EAT–*Lancet* reference diet and mortality, stroke, and greenhouse gas emissions in three cohorts. *Lancet Planet*. Health 8, e391–e401 (2024).

22. Satija, A. & Hu, F. B. Plant-based diets and cardiovascular health. Trends Cardiovasc. Med. 28, 437–441 (2018).

23. Abu-Saad, K. et al. Adaptation and predictive utility of a Mediterranean diet screener score. Clin. Nutr. 38, 2928–2935 (2019).

25. Ke, G. et al. LightGBM: a highly efficient gradient boosting decision tree. in *Proceedings of the 31st International Conference on Neural Information Processing Systems* 3149–3157 (Curran Associates Inc., Red Hook, NY, USA, 2017).

26. de la Cuesta-Zuluaga, J. et al. Age- and Sex-Dependent Patterns of Gut Microbial Diversity in Human Adults. mSystems 4, 10.1128/msystems.00261-19 (2019).

27. Lundberg, S. M. et al. From local explanations to global understanding with explainable AI for trees. *Nat*. Mach. Intell. 2, 56–67 (2020).

28. Belda, I. et al. A multi-omics approach for understanding the effects of moderate wine consumption on human intestinal health. Food Funct. 12, 4152–4164 (2021).

29. Benjamini, Y. & Hochberg, Y. Controlling the False Discovery Rate: A Practical and Powerful Approach to Multiple Testing. J. R. Stat. Soc. Ser. B Methodol. 57, 289–300 (1995).

30. Manghi, P. et al. Coffee consumption is associated with intestinal Lawsonibacter asaccharolyticus abundance and prevalence across multiple cohorts. Nat. Microbiol. 9, 3120–3134 (2024).

31. Fijan, S. Microorganisms with Claimed Probiotic Properties: An Overview of Recent Literature. Int. J. Environ. Res. Public. Health 11, 4745–4767 (2014).

32. Aslam, H. et al. The effects of dairy and dairy derivatives on the gut microbiota: a systematic literature review. Gut Microbes 12, 1799533.

33. JanssenDuijghuijsen, L. et al. Changes in gut microbiota and lactose intolerance symptoms before and after daily lactose supplementation in individuals with the lactase nonpersistent genotype. Am. J. Clin. Nutr. 119, 702–710 (2024).

34. Cooper, D. N., Martin, R. J. & Keim, N. L. Does Whole Grain Consumption Alter Gut Microbiota and Satiety? Healthcare 3, 364–392 (2015).

35. Mano, F. et al. The Effect of White Rice and White Bread as Staple Foods on Gut Microbiota and Host Metabolism. Nutrients 10, 1323 (2018).

36. Ben-Yacov, O. et al. Gut microbiome modulates the effects of a personalised postprandial-targeting (PPT) diet on cardiometabolic markers: a diet intervention in pre-diabetes. Gut 72, 1486–1496 (2023).

37. Carlino, N. et al. Unexplored microbial diversity from 2,500 food metagenomes and links with the human microbiome. Cell 187, 5775–5795.e15 (2024).

38. Beghini, F. et al. Integrating taxonomic, functional, and strain-level profiling of diverse microbial communities with bioBakery 3. eLife 10, e65088 (2021).

39. Fernández-Bautista, T., Gómez-Gómez, B., Gracia-Lor, E., Pérez-Corona, T. & Madrid, Y. Investigating the Presence of Selenoneine, Ergothioneine, and Selenium-Containing Biomolecules in Fish and Fish-Derived Commercial Products. J. Agric. Food Chem. 72, 26155–26164 (2024).

40. Tan, J. et al. Chapter Three - The Role of Short-Chain Fatty Acids in Health and Disease. in Advances in Immunology (ed. Alt, F. W.) vol. 121 91–119 (Academic Press, 2014).

41. Hamer, H. M. et al. Review article: the role of butyrate on colonic function. Aliment. Pharmacol. Ther. 27, 104–119 (2008).

42. Zhou, T. et al. High-resistant starch and low-glutelin content 1 rice benefits gut function in obese patients. Front. Sustain. Food Syst. 8, (2024).

43. Martinez Tuppia, C. et al. In Vitro Human Gastrointestinal Digestibility and Colonic Fermentation of Wheat Sourdough and Yeast Breads. Foods 13, 3014 (2024).

44. Larsen, P. E. & Dai, Y. Modeling interaction networks between host, diet, and bacteria predicts obesogenesis in a mouse model. Front. Mol. Biosci. 9, (2022).

45. Joos, R. et al. Examining the healthy human microbiome concept. Nat. Rev. Microbiol. 23, 192–205 (2025).

46. Xia, M. et al. Structural and functional alteration of the gut microbiota in elderly patients with hyperlipidemia. Front. Cell. Infect. Microbiol. 14, (2024).

47. Bancil, A. S. et al. Food Additive Emulsifiers and Their Impact on Gut Microbiome, Permeability, and Inflammation: Mechanistic Insights in Inflammatory Bowel Disease. J. Crohns Colitis 15, 1068–1079 (2021).

48. Naimi, S., Viennois, E., Gewirtz, A. T. & Chassaing, B. Direct impact of commonly used dietary emulsifiers on human gut microbiota. Microbiome 9, 66 (2021).

49. Asnicar, F. et al. Microbiome connections with host metabolism and habitual diet from 1,098 deeply phenotyped individuals. Nat. Med. 27, 321–332 (2021).

50. Taylor, B. C. et al. Consumption of Fermented Foods Is Associated with Systematic Differences in the Gut Microbiome and Metabolome. mSystems 5, e00901–19 (2020).

51. Leeuwendaal, N. K., Stanton, C., O’Toole, P. W. & Beresford, T. P. Fermented Foods, Health and the Gut Microbiome. Nutrients 14, 1527 (2022).

52. Wastyk, H. C. et al. Gut-microbiota-targeted diets modulate human immune status. Cell 184, 4137–4153.e14 (2021).

53. Berglund, L. et al. Evaluation and Treatment of Hypertriglyceridemia: An Endocrine Society Clinical Practice Guideline. J. Clin. Endocrinol. Metab. 97, 2969–2989 (2012).

54. Castro-Quezada, I. et al. Glycemic Index, Glycemic Load and Dyslipidemia in Adolescents from Chiapas, Mexico. Nutrients 16, 1483 (2024).

55. Luna-Castillo, K. P. et al. The Effect of Dietary Interventions on Hypertriglyceridemia: From Public Health to Molecular Nutrition Evidence. Nutrients 14, 1104 (2022).

56. Leviatan, S., Shoer, S., Rothschild, D., Gorodetski, M. & Segal, E. An expanded reference map of the human gut microbiome reveals hundreds of previously unknown species. Nat. Commun. 13, 3863 (2022).

57. Pedregosa, F. et al. Scikit-learn: Machine Learning in Python. J Mach Learn Res 12, 2825–2830 (2011).

58. Blanco-Míguez, A. et al. Extending and improving metagenomic taxonomic profiling with uncharacterized species using MetaPhlAn 4. Nat. Biotechnol. 41, 1633–1644 (2023).

