## Extended Figures for "Diet shapes the gut microbiome: cross-sectional and longitudinal insights from the Human Phenotype Project"

### Extended Data Fig. 1: Extended diversity and abundance predictions.

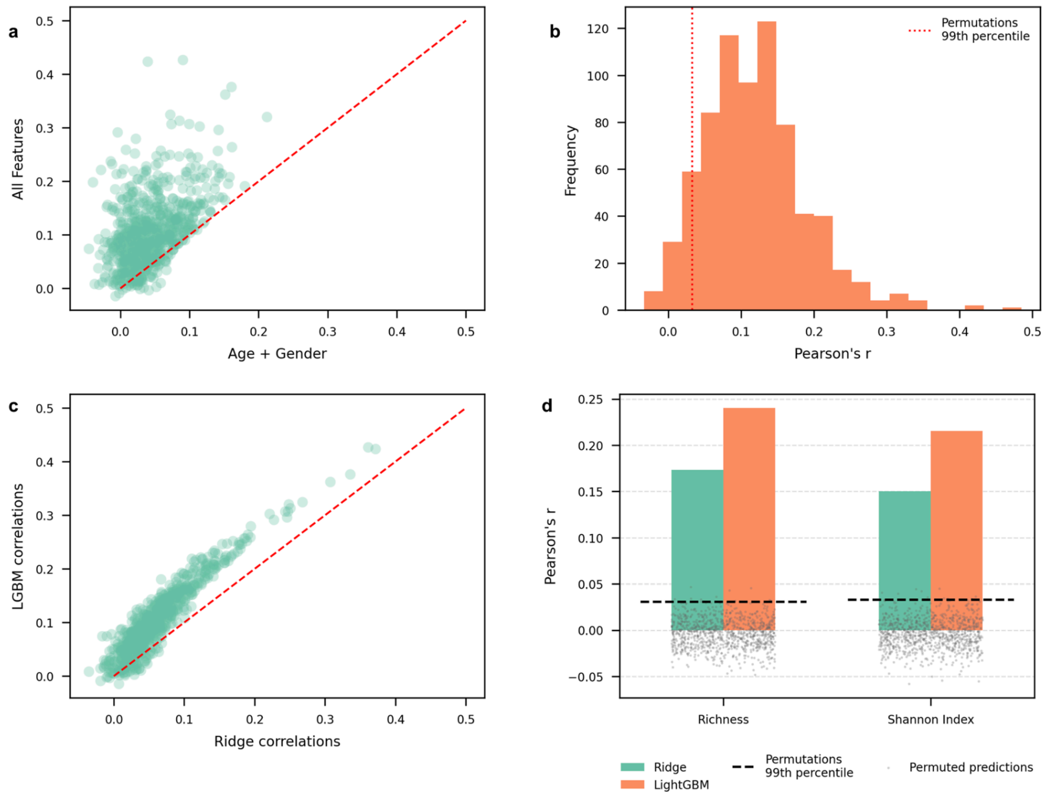


**a.** Comparison of species abundance prediction performance between models trained with age and sex only (x-axis) and with all features (y-axis). Each dot represents a species; the red dashed line indicates the identity line. **b.** Histogram of Pearson correlations (LGBM) between predicted and observed species abundances, in the test set. A red line indicates the 99th percentile of correlations obtained from permutation testing, highlighting significant correlations. **c.** Comparison of species abundance prediction performance between ridge regression (x-axis) and LightGBM (y-axis). Each dot represents a species; the red dashed line indicates the identity line. **d.** Diversity targets linear model versus LGBM model. Pearson correlation coefficients (r) for ridge linear regression models (blue) and LightGBM models (orange) trained to predict species richness and Shannon diversity. Grey dots represent permuted Pearson's r with all features, and dashed line indicates 99th percentile of permutations.

### Extended Data Fig. 2: SHAP summary plots of highly predicted microbial species.

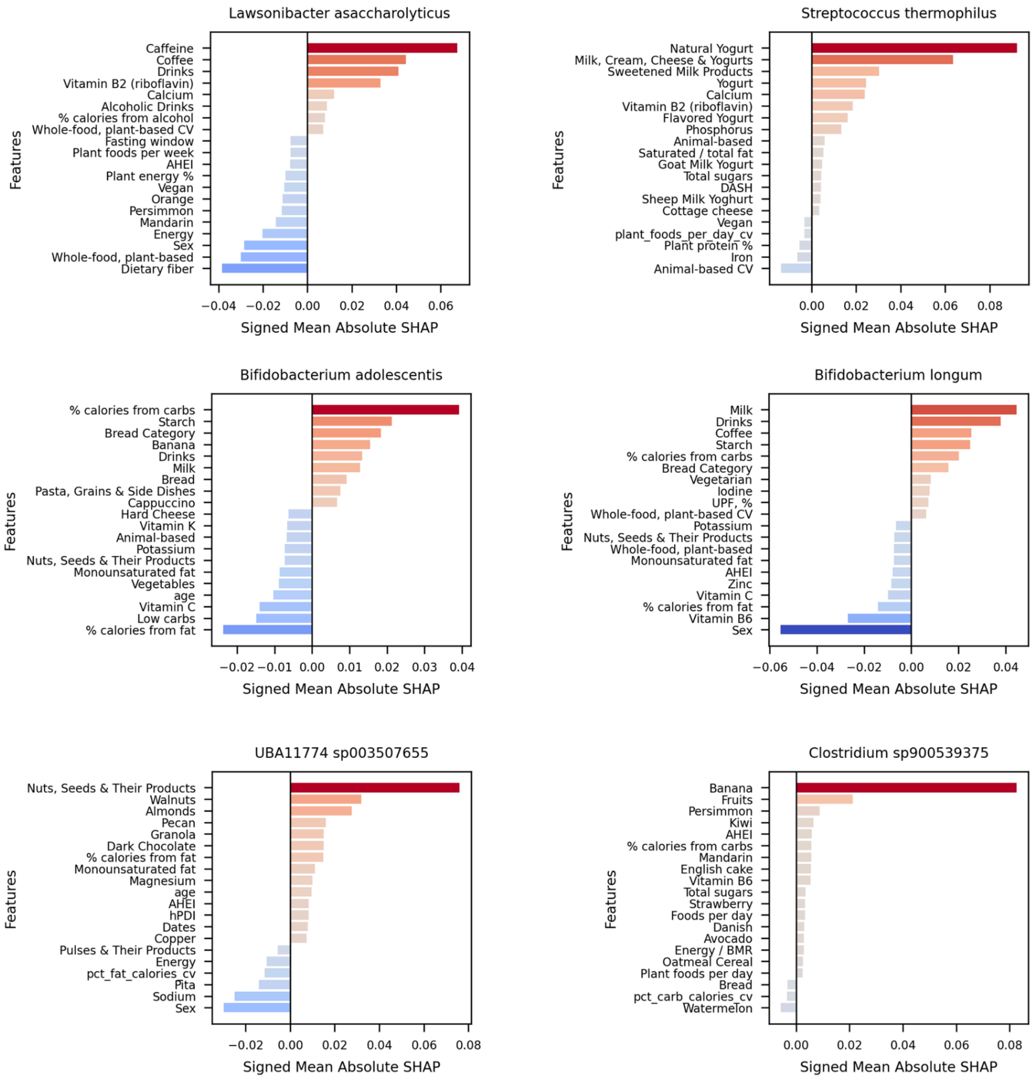


SHAP summary plot for highly predicted species, shows feature contributions sorted by importance; SHAP direction was derived from spearman correlation directions.

**Extended Data Fig. 3: Presence of diet-predictive microbial species in foods from the curated Food Metagenomic Database (cFMD).**

a

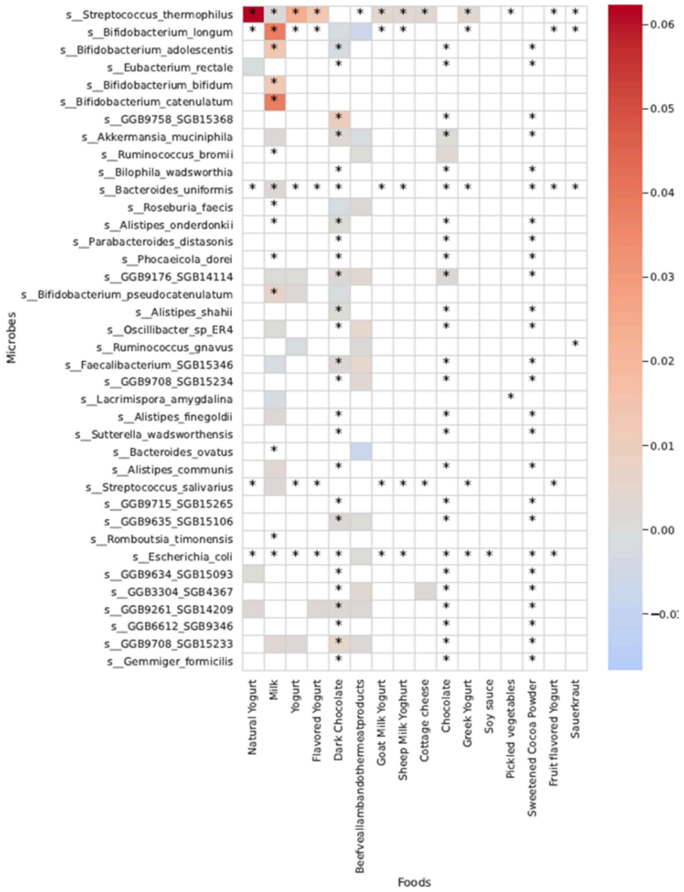


**a.** A heatmap showing microbial species detected across 16 food items (columns) and 38 species with significant predictive associations in the cohort (rows). Colors indicate mean relative abundance (MetaPhlAn-derived), with warmer shades reflecting higher abundance. Asterisks mark species-food pairs where the species was detected (abundance > 0). Several key associations were confirmed: Streptococcus thermophilus was abundant in yogurt, consistent with its role in fermentation; milk contained Bifidobacterium longum and Bifidobacterium adolescentis; and UBA11774 sp003507655, primarily predicted by nut intake, was detected in dark chocolate. Conversely, some strong dietary predictors (e.g., Lawsonibacter asaccharolyticus for coffee) were not found in food sequencing.

**Extended Data Fig. 4: Microbiome functional pathways are highly predicted by diet.**

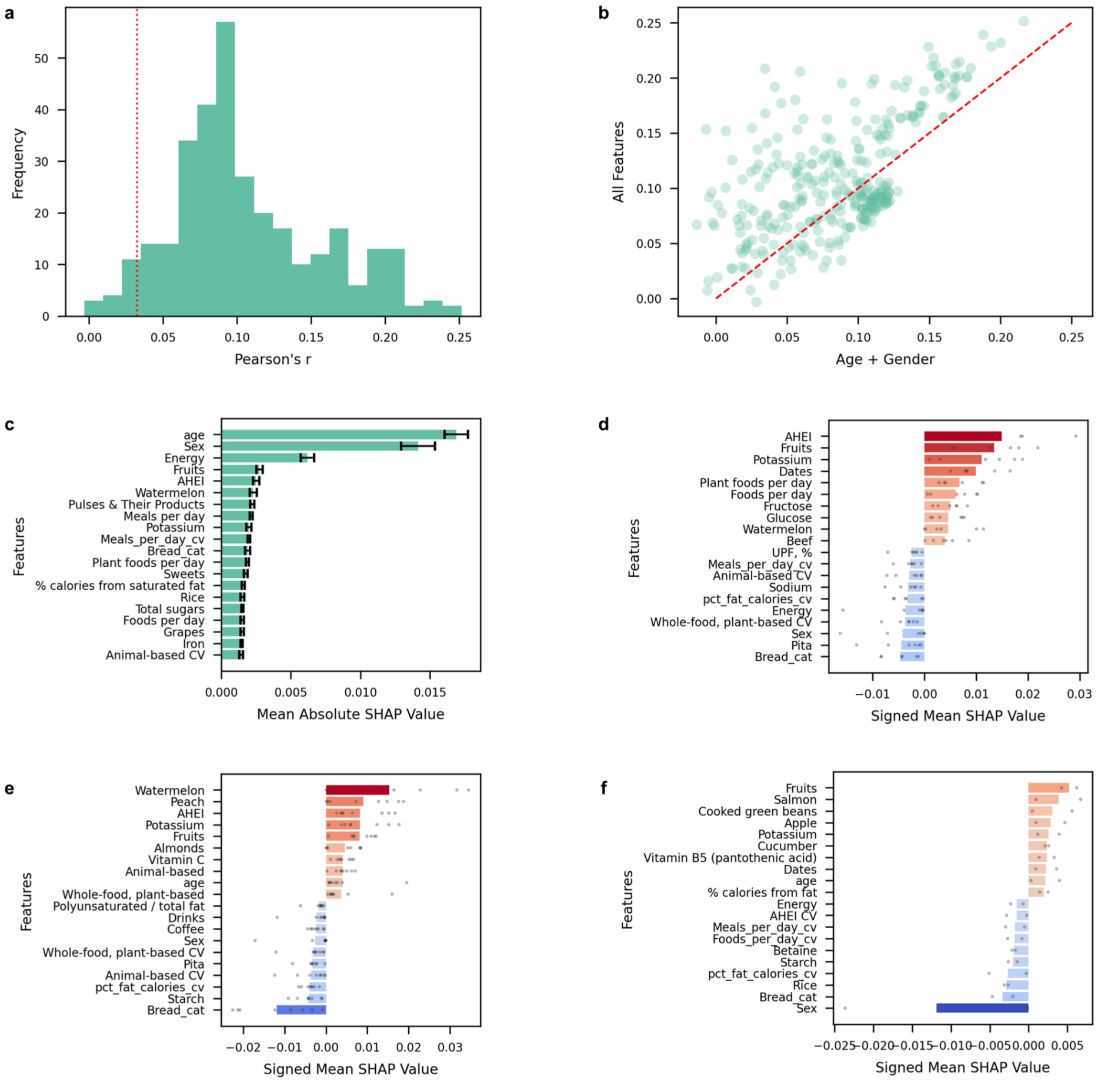


**a.** Histogram of Pearson correlations (LGBM) between predicted and observed pathway abundances. A red line indicates the 99th percentile of correlations obtained from permutation testing, highlighting significant correlations. **b.** Pearson correlations of pathway predictions from baseline models (age+sex) compared with models including all features. **c.** Mean absolute SHAP values averaged across all pathways; error bars show the standard error of the mean across species. Higher values reflect greater average importance for prediction. **d-f.** Directional SHAP values for representative functional pathway groups. Grey dots denote individual pathways, bars show mean signed SHAP values. d, Biosynthetic pathways. e, Carbohydrate and plant polysaccharide degradation pathways. f, Butyrate producing pathways.

**Extended Data Fig. 5: Main predictors heatmap of SHAP values for the most food-predictable pathways.**

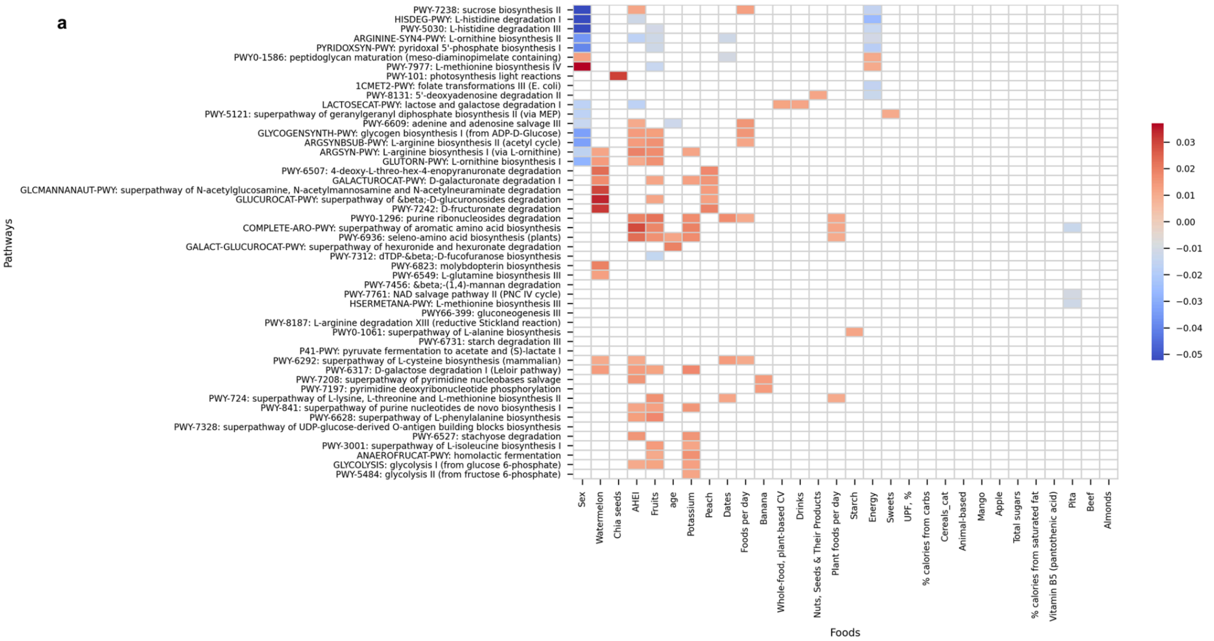


**a.** Main predictors heatmap of SHAP values for the top 50 pathways with highest predictive gain from baseline, and the 30 dietary features with the highest maximal importance. We clustered species by pathways in their SHAP value profiles across foods. SHAP values < 0.01 were masked.

### Extended Data Fig. 6: Predictive gain from baseline of various health phenotypes from gut microbiome.

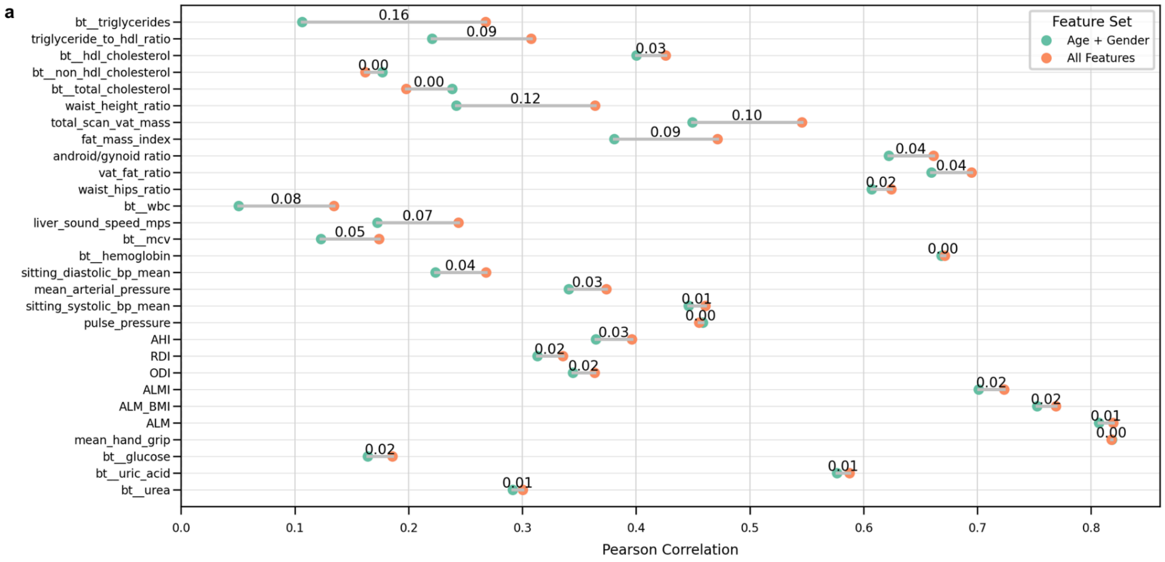


**a.** Pearson correlations between predicted and observed values for clinically relevant health phenotypes. For each phenotype, circles show predictive performance of a baseline model including age and sex (green) and a microbiome-informed model including species abundances in addition to age and sex (orange). Lines connect the paired values for each phenotype, with numbers indicating the improvement in correlation (Δr). Five-fold cross-validation was used to generate predictions. All phenotypes shown were significantly predicted by the microbiome-informed model (FDR < 0.05).

**Extended Data Fig. 7: Personalized dietary simulated interventions predict microbiome shifts and VAT reductions.**

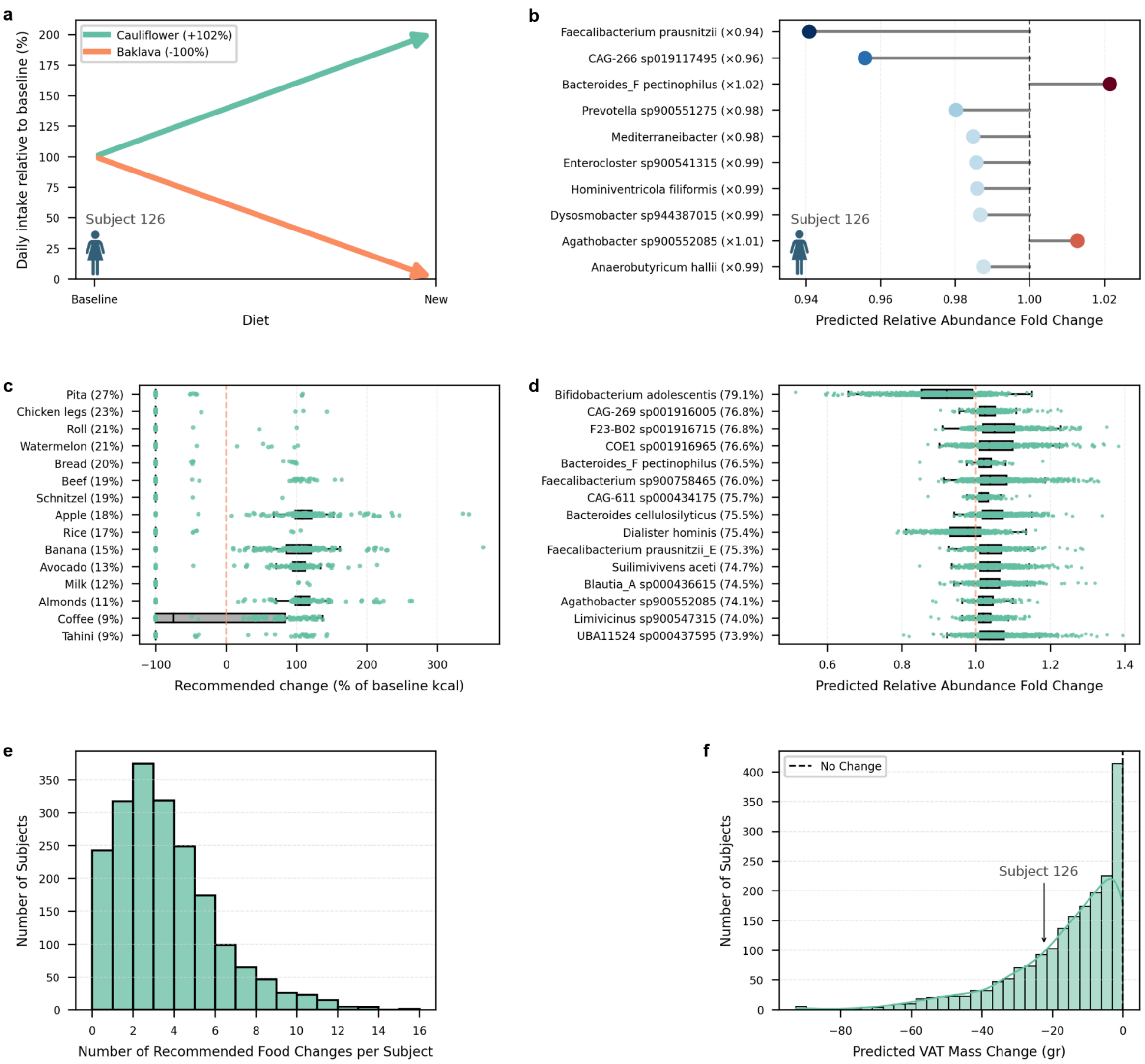


**a.** Personalized dietary recommendations for a representative individual (61-65 years old female). Arrows indicate foods predicted to increase or decrease relative to baseline intake, with percent changes shown in the legend. **b.** Predicted microbial shifts for the same individual, showing the ten species with the largest absolute fold changes. **c.** Most frequent food changes recommended across the cohort. Percentages show how many individuals who ate each food were advised to change it. Recommended changes are given as percent of baseline calories; each point is one individual. **d.** Microbial species most frequently predicted to change in response to the recommended diets. Percentages show how many individuals carrying each species were predicted to see a change. **e.** Number of recommended food changes per individual. Most people received only a few changes, with a median of three. **f.** Predicted changes in visceral adipose tissue (VAT) mass across individuals. The dashed line marks no change, and the arrow highlights a predicted 22 gr decrease in VAT mass for the representative individual shown in panels a and b.

### Extended Data Fig. 8: Personalized dietary simulated interventions predict microbiome shifts and WBC reductions.

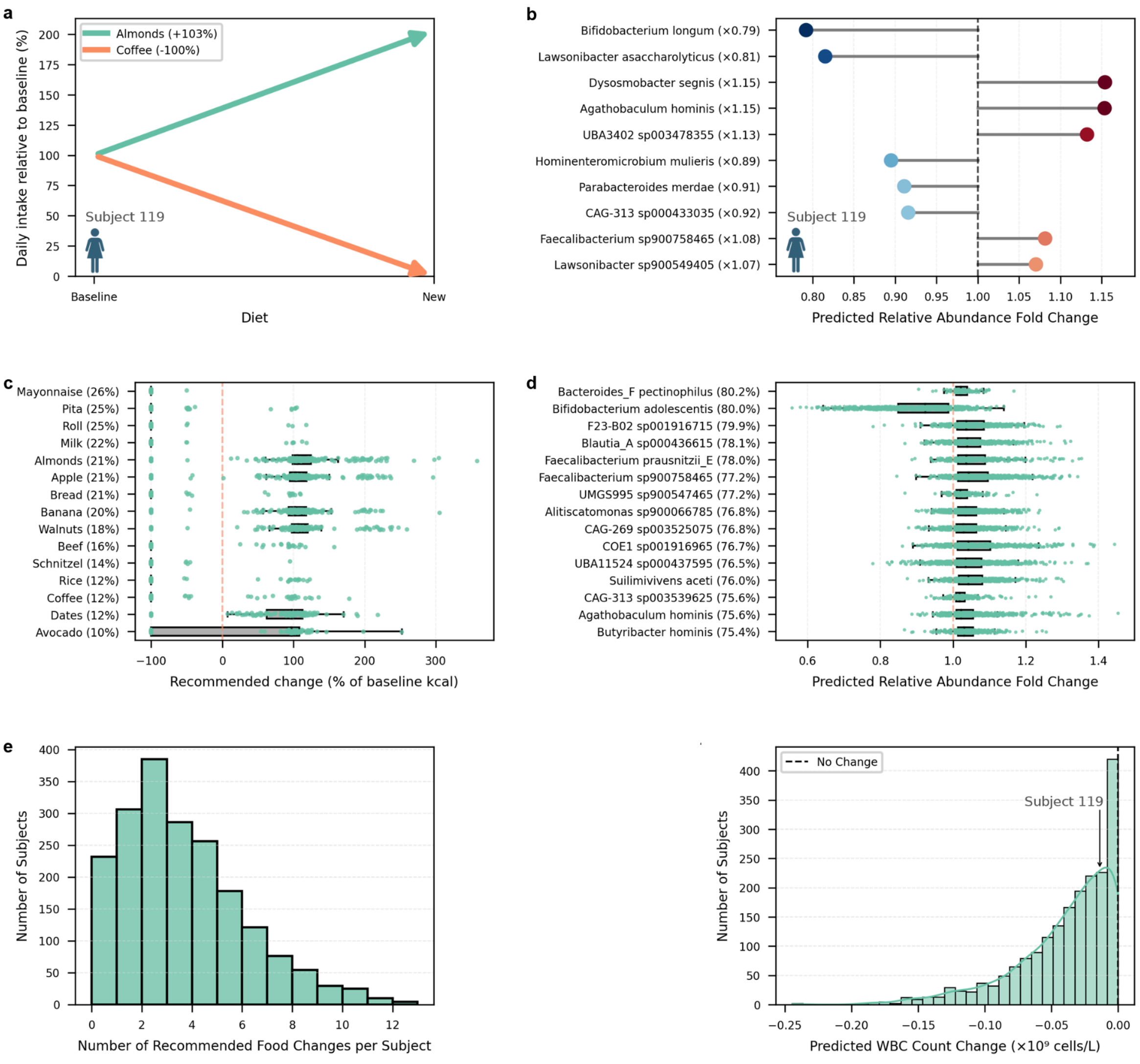


**a.** Personalized dietary recommendations for a representative individual (56-60 years old female). Arrows indicate foods predicted to increase or decrease relative to baseline intake, with percent changes shown in the legend. **b.** Predicted microbial shifts for the same individual, showing the ten species with the largest absolute fold changes. **c.** Most frequent food changes recommended across the cohort. Percentages show how many individuals who ate each food were advised to change it. Recommended changes are given as percent of baseline calories; each point is one individual. **d.** Microbial species most frequently predicted to change in response to the recommended diets. Percentages show how many individuals carrying each species were predicted to see a change. **e.** Number of recommended food changes per individual. Most people received only a few changes, with a median of three. **f.** Predicted changes in white blood cell (WBC) count across individuals. The dashed line marks no change, and the arrow highlights a predicted decrease of $0.014\times10^{9}$ cells/L in WBC count for the representative individual shown in panels a and b.
